# Financial and Non-financial Conflicts of Interest Among the Japanese Government Advisory Board Members Concerning Coronavirus Disease 2019

**DOI:** 10.1101/2021.09.12.21263442

**Authors:** Hanano Mamada, Anju Murayama, Akihiko Ozaki, Takanao Hashimoto, Hiroaki Saito, Toyoaki Sawano, Divya Bhandari, Sunil Shrestha, Tetsuya Tanimoto

**Affiliations:** Medical Governance Research Institute, Minato-ku, Tokyo, Japan; Akita University School of Medicine, Akita, Akita, Japan; Tohoku University School of Medicine, Sendai, Miyagi, Japan; Department of Breast Surgery, Jyoban Hospital of Tokiwa Foundation, Iwaki, Fukushima, Japan; Department of Pharmacy, Sendai City Medical Center, Sendai, Miyagi, Japan; Department of Gastroenterology, Sendai Kousei Hospital, Sendai, Miyagi, Japan; Department of Surgery, Jyoban Hospital of Tokiwa Foundation, Iwaki, Fukushima, Japan; School of Pharmacy, Monash University Malaysia, Jalan Lagoon Selatan, Bandar Sunway, Selangor Darul Ehsan, Malaysia; Department of Internal Medicine, Navitas Clinic, Kawasaki, Kanagawa, Japan

**Author notes:** **Corresponding author** Anju Murayama, Medical Governance Research Institute, Minato-ku, Tokyo, 1087505, Japan. The authors equally contributed to this study (Hanano Mamada, Anju Murayama).

**Keywords:** Conflict of interest, COVID-19, advisory board, ethics, Japan, coronavirus disease 2019, SARS-CoV-2

## Abstract

**Objectives:** This study aimed to assess the extent of conflicts of interest among the Japanese government COVID-19 advisory board members and elucidate the accuracy of conflicts of interest (COI) disclosure and management strategies.

**Methods:** Using the payment data from all 79 pharmaceutical companies in Japan between 2017 and 2018 and direct research grants from the Japanese government between 2019 and 2020, we evaluated the extent of financial and non-financial COI among all 20 Japanese government COVID-19 advisory board members.

**Results:** Japanese government COVID-19 advisory board members were predominantly male (75.0%) and physicians (50.0%). Between 2019 and 2020, two members (10.0%) received a total of $819,244 in government research funding. Another five members (25.0%) received $419,725 in payments, including $223,183 in personal fees, from 28 pharmaceutical companies between 2017 and 2018. The average value of the pharmaceutical payments was $20,986 (standard deviation: $81,762). Further, neither the Ministry of Health, Labor, and Welfare nor the Japanese Cabinet Secretariat disclosed financial or non-financial COI with industry. Further, the government and had no policies for managing COI among advisory board members.

**Conclusions:** This study found that the Japanese government COVID-19 advisory board had financial and non-financial COI with pharmaceutical companies and the government. Further, there were no rigorous COI management strategies for the COVID-19 advisory board members. Any government must ensure the independence of scientific advisory boards by implementing more rigorous and transparent management strategies that require the declaration and public disclosure of all COI.

## Introduction

The United States (U.S.) National Academy of Medicine defines conflicts of interest (COI) as “circumstances that create a risk that professional judgments or actions regarding a primary interest will be unduly influenced by a secondary interest.”[1] Both financial and non-financial COI exist.[1] According to a U.S. Preventive Services Task Force review, non-financial COI include competing beliefs, public comments, interests, advocacy, or policy positions, substantial career efforts, or intellectual ones. Intellectual COI include study authorship or grant funding directly related to the topic that creates the potential for bias and compromises objectivity in judgment.[2] Given the nature of non-financial COI, they could be particularly prominent individuals in positions close to public authorities. Consequently, non-financial COI may be even more influential than financial ones.[2, 3] While financial COI garner a great deal of attention over transparency and financial thresholds,[4] non-financial COI receive little attention, even among national health policy experts.[5]

This lack of discussion over non-financial COI has important implications for the ongoing coronavirus 2019 (COVID-19) pandemic. As of June 24, 2021, more than 3.8 million people have died from COVID-19.[6] Increasingly, accumulating evidence from developing and developed countries suggests the potential to minimize the damage of diseases like COVID-19 by appropriately assigning competent and fair-minded experts to government advisory boards. For governments to gain unbiased, evidence-based advice from medical and public health experts and implement evidence-based health policies relating to COVID-19, proper management of these experts COI is essential in any country.[4, 7]

Unfortunately, the Japanese government lags behind other countries in several COVID-19 prevention strategies, including severe acute respiratory syndrome coronavirus 2 (SARS-CoV-2) infection control and vaccination efforts.[8] Japan has also received global criticism for hosting the Olympic Games despite poorly developed strategies for managing COVID-19.[9, 10] Therefore, it remains vital to investigate the backgrounds of the experts who developed the COVID-19 policies in Japan and how they managed their COI. This study aimed to evaluate the extent and prevalence of financial and non-financial COI between pharmaceutical companies and the Japanese government and elucidate the accuracy of disclosure and management strategies for COI among the COVID-19 advisory board members.

## Methods

In response to the domestic spread of SARS-CoV2, on February 24, 2020, the Japanese government established the Expert Meeting on Novel Coronavirus Disease Control Committee (EMNCDCC) under the Ministry of Health, Labor, and Welfare. This committee consisted of infectious disease, microbiology, and public health experts, charged with developing strategies to prevent COVID-19 transmission. A subsequent reorganization followed in July 2020 to tackle this infectious disease from a more multidimensional perspective. The changes included expanding the committee to include economists, managers, and jurists, rebranding as the Subcommittee on Novel Coronavirus Disease Control (SNCDC), and reassignment under the Cabinet Secretariat. We identified the current SNCDC composition, as of June 1, 2021, from the Japanese Cabinet Secretariat website.[11]

We then collected demographic data on the most recent SNCDC members, including gender, position, and specialty, from official documents and webpage of affiliated institutions. The gender of the board members was verified by the database of medical physician published by the Ministry of Health, Labor and Welfare for the board members with a Japanese medical license (https://licenseif.mhlw.go.jp/search_isei/jsp/top.jsp), the photographs on the official page of affiliated institutions after a manual Google search, as well as news articles such as interview with the board members in the major national newspapers. We extracted the h-index for researchers and physicians using the Scopus database (https://www.scopus.com/freelookup/form/author.uri), or when not accessible, Google Scholar to evaluate the academic performance of individual SNCDC members, as described previously.[12] For financial COI, we examined payment records from pharmaceutical companies.

Payment data included speaking fees, writing fees, consulting fees, and scholarship donations disclosed by 79 Japanese pharmaceutical companies between 2017 and 2018. Most international professional and academic associations consider the three prior years as an appropriate period for COI disclosure.[2] The latest available payment data for analysis in Japan came from 2018, as in our previous studies.[13, 14] For research funding from the government, we used the Research Program on Emerging and Re-emerging Infectious Diseases in 2019 (https://www.mhlw.go.jp/seisakunitsuite/bunya/hokabunya/kenkyujigyou/hojokin-koubo-2019/gaiyo/16.html) and 2020 (https://www.mhlw.go.jp/seisakunitsuite/bunya/hokabunya/kenkyujigyou/hojokin-koubo-2020/gaiyo/16.html). This program dispensed and publicly disclosed funding awards from the Ministry of Health, Labor, and Welfare and was considered as potential non-financial COI with the government.

We conducted descriptive analysis for both financial and non-financial COI among the SNCDC members with the government and industry. Japanese yen were converted to U.S. dollars (USD) using a yearly average monthly exchange rate of 112.1, 110.4, 109.0, and 106.8 per USD for 2017, 2018, 2019, and 2020. Further, when we could not find information on COI disclosure among the SNCDC members, we contacted the Japanese Cabinet Secretariat and the Ministry of Health, Labor, and Welfare about their disclosure and management strategies for the EMNCDCC and SNCDC by telephone. The Ethics Committee of the Medical Governance Research Institute approved this study on June 5, 2020. (ID: MG2018-04-0516) Informed consent waived and direct contact to the related organizations, including the Japanese Cabinet Secretariat and the Japanese Ministry of Health, Labor, and Welfare, were allowed by the Ethics Committee of the Medical Governance Research Institute.

## Results

As of June 1, 2021, the 20 identified current SNCDC members were predominantly male (75.0%), physicians (50.0%), and had a median h-index of 18.5 (Interquartile ranges: 4.5-32) (Table 1). Additionally, two (10.0%) received a total of $819,244 in research payments from the Ministry of Health, Labor, and Welfare between 2019 and 2020, while five (25.0%) received $419,724 in payments that included $223,182 in personal payments and $196,543 as scholarships from 33 pharmaceutical companies between 2017 and 2018 (Table 2). The average (standard deviation, SD) pharmaceutical company payments made to the 20 advisory board members were $20,986 ($81,762). Of the 28 pharmaceutical companies making payments to the advisory board members, FujiFilm Toyama Chemical Co., Ltd. ($86,657) made the largest individual payments, followed by Daiichi Sankyo ($62,968) and Pfizer ($61,871).

**Table 1.**
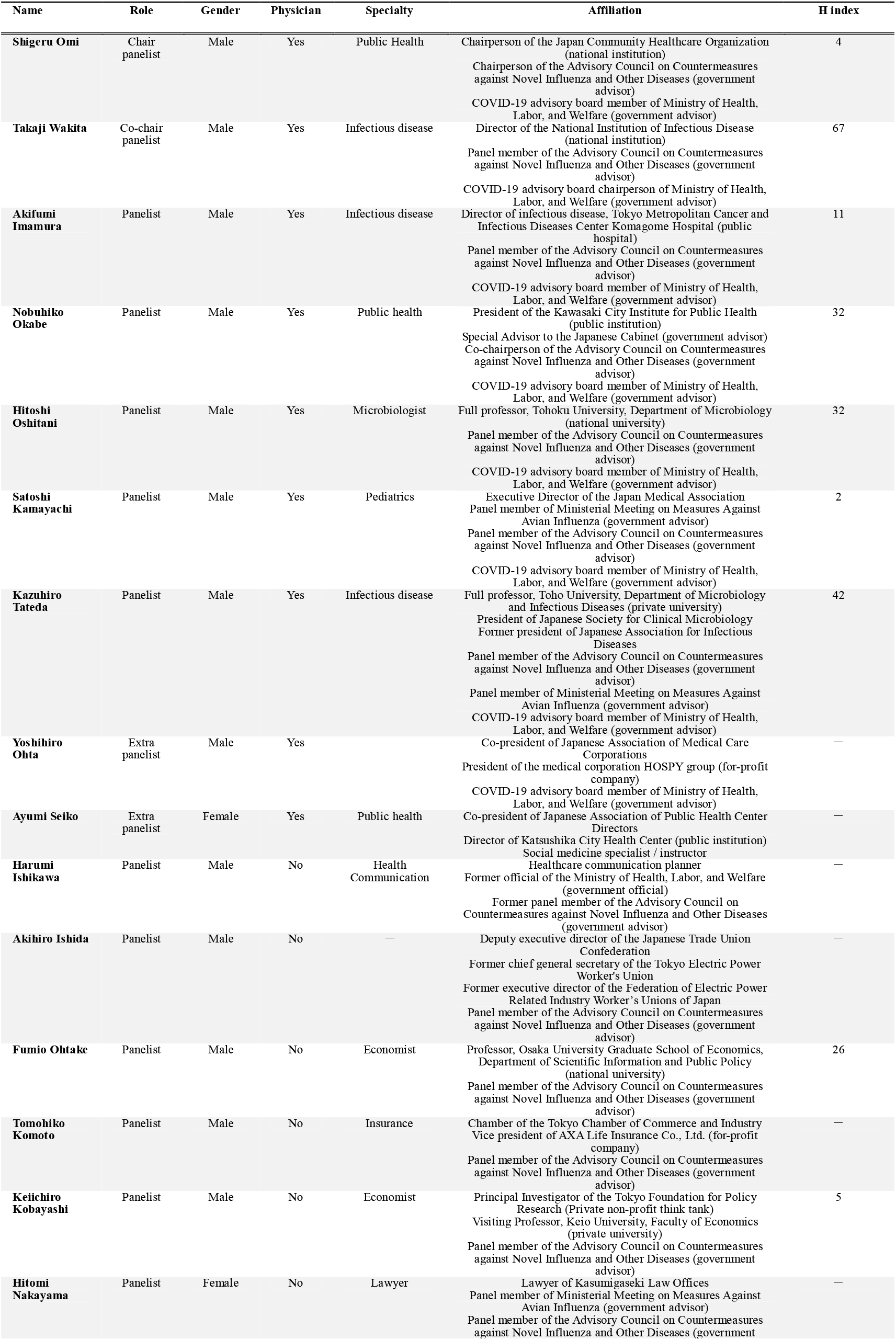
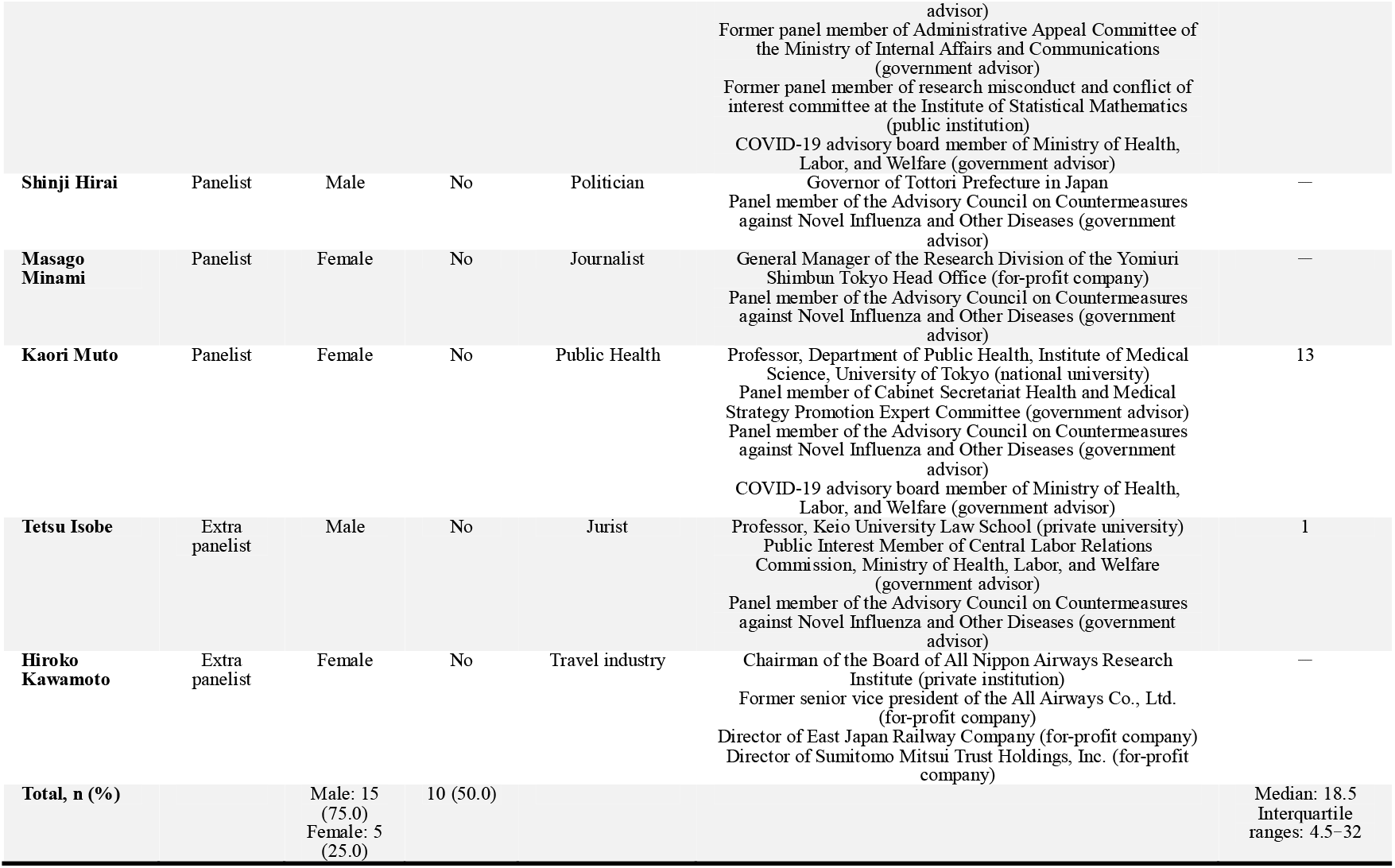
Panel member list of the Subcommittee on Novel Coronavirus Disease Control.

**Table 2.** **Payments to the Subcommittee on Novel Coronavirus Disease Control panel members from pharmaceutical companies between 2017 and 2018 and from the Ministry of Health, Labor, and Welfare between 2019 and 2020**

| Name | Pharmaceutical company payments (\$) | | | Japanese government payments (\$) | | Total Received (\$) |
| --- | --- | --- | --- | --- | --- | --- |
|  | Personal payments | Scholarship donations | Total | In 2019 | In 2020 |  |
| **Akifumi Imamura** | 8,939 | 0 | 8,939 | 0 | 0 | 8,939 |
| **Nobuhiko Okabe** | 16,723 | 0 | 16,723 | 287,156 | 252,125 | 556,004 |
| **Hitoshi Oshitani** | 0 | 0 | 0 | 0 | 0 | 0 |
| **Satoshi Kamayachi** | 0 | 0 | 0 | 0 | 0 | 0 |
| **Kazuhiro Tateda** | 180,188 | 196,543 | 376,730 | 0 | 0 | 376,730 |
| **Kaori Muto** | 2,598 | 0 | 2,598 | 0 | 279,963 | 282,561 |
| **Yoshihiro Ohta** | 14,734 | 0 | 14,734 | 0 | 0 | 14,734 |
| **Total** | 223,182 | 196,543 | 419,724 | 287,156 | 532,088 |  |

Surprisingly, we found that neither the Ministry of Health, Labor, and Welfare, nor the Japanese Cabinet Secretariat, disclosed the financial and non-financial COI of SNCDC members. Following our inquiry about COI, an official representative of the Japanese Cabinet Secretariat indicated they did not ask the SNCDC members to declare their COI. Additionally, SNCDC members were selected from an existing government advisory board, the Advisory Council on Countermeasures against Novel Influenza and Other Diseases. Similarly, the Ministry of Health, Labor, and Welfare did not ask the members of the EMNCDCC to declare their COI. Thus, members of either committee were not required to declare or disclose their COI.

## Discussion

We found substantial financial and non-financial COI among the COVID-19 advisory board members with pharmaceutical companies and the Japanese government itself. Further, no COI policies or management strategies exist to ensure disclosure of COI by the COVID-19 advisory board members.

With the recent global demand for greater transparency and growing evidence that financial and non-financial COI can influence professional judgment and behaviors,[15–19] there remains a strong need for integrity among healthcare professionals. Current trends in developed nations favor the full public disclosure of all financial and non-financial COI, coupled with rigorous and appropriate management, in building and maintaining public trust.[20–23] The first step in this process requires improved transparency, which inherently relies on full COI declaration and disclosure by healthcare professionals. Indeed, for this reason, several other government COVID-19 advisory boards require members to declare COI for public disclosure. Such organizations include the United Kingdom’s (U.K.) Scientific Advisory Group for Emergencies[24] and the Joint Committee on Vaccination and Immunization[4, 25], the Scottish Government’s Covid-19 Advisory Group,[26] the COVID-19 Immunity Task Force in Canada,[27] and in the U.S. the Advisory Committee on Immunization Practice in Centers for Disease Control and Prevention[28, 29] and the COVID-19 authorization committees at the U.S. Food and Drug Administration.[4] Unfortunately, despite most SNCDC members having close ties to the Japanese government, whether from federal research funding, current employment, or past or present service on government committees, SNCDC members were not required to, nor have they voluntarily declared or disclosed their COI even a year after the onset of the COVID-19 pandemic. This poses a potentially serious problem for COI management. Suppose the public and outside experts do not have access to the expert decision-making processes of governments or government advisory bodies on the COVID-19 pandemic. In this case, they can never evaluate previous policies and prepare for future pandemics. Thus, the Japanese government should urgently implement appropriate COI management strategies centered on greater transparency, ensuring the immediate and complete disclosure of SNCDC members’ COI.

Notably, we found that two SNCDC members received more than $250,000 in research grants from the Ministry of Health, Labor, and Welfare for studies on COVID-19 in 2020. Funding of this sort, straight from the government for research directly related to SNCDC functions, represents non-financial COI as recently defined by the U.S. Preventive Services Task Force and other international organizations.[2]

Such direct non-financial COI between the government and advisory board members may inappropriately influence future recommendations. Instead, government experts should manage their COI appropriately and make recommendations based solely on scientific evidence, uninfluenced by their non-financial COI with the government. This is critical as subject matter experts should act and frame their opinions based on evidence in tackling the pandemic on behalf of the public and preventing further damage to human lives. Unfortunately, no formal definition of non-financial COI or strategies addressing COI management exist in Japan. Although the influence of non-financial COI among the government advisory board members on their actions and recommendations is beyond our study setting, all such panelists in any country should recognize how COI might influence their behavior and manage them appropriately.[19]

As for financial COI, five SNCDC members received payments from pharmaceutical companies developing or manufacturing COVID-19 drugs or vaccines, including Pfizer, whose vaccines were the first approved in Japan. Other companies making payments to SNCDC members include Daiichi Sankyo, whose mRNA COVID-19 vaccine is currently in Phase □/□ clinical trials, and FujiFilm, whose potential drug for treating COVID-19, favipiravir (Avigan), is in phase □ trial sponsored by the Ministry of Health, Labor and Welfare. Also, among the five SNCDC members receiving personal payments from pharmaceutical companies, three received more than $10,000, and one received $180,188 between 2017 and 2018. Additionally, in 2017, one panelist received $118,644 in scholarship donations from 10 pharmaceutical companies, including $53,798 from FujiFilm; $47,554 from Daiichi Sankyo, the manufacturer of the COVID-19 vaccine developed by AstraZeneca in Japan; and $18,884 from Shionogi whose COVID-19 vaccine is currently in Phase □/□.

The ongoing debate over financial COI among government advisory board members extends beyond Japan. It includes other developed nations like the U.K. and the U.S.[4] As an investigative journalist, Thacker (2020) reported that members of the Scientific Advisory Group for Emergencies were required to declare their financial COI. However, the U.K. government failed to disclose this information to the British Medical Journal in 2020.[7] Additionally, Thacker (2021) found that several members of the U.K.’s Joint Committee on Vaccines and Immunization had undisclosed financial relationships with pharmaceutical companies, and similarly, that members of the U.S. Food and Drug Administration had received substantial sums from pharmaceutical companies.[4] Given that most international healthcare organizations and professional organizations set the current reporting periods for financial COI declaration,[2, 30] undoubtedly the receipt of payments from the pharmaceutical companies developing or manufacturing COVID-19 drugs and vaccines represent financial COI. Such information must be managed and disclosed to the public. However, the EMNCDCC and SNCDC members were not required to declare their COI even to the government.

Unfortunately, both the EMNCDCC and SNCDC members and the Japanese government made several mistakes concerning the COVID-19 pandemic. First, the government and both the EMNCDCC and SNCDC members emphasized controlling clusters, relying heavily on behavior modification, and preventing infections in enclosed or crowded spaces and close contact environments, collectively known as the “3Cs.”[31] However, these strategies failed to prevent further transmission in the presence of large numbers of COVID-19 cases with unknown transmission routes.[32] Indeed, the Japanese government declared and extended a state emergency several times in response to mounting case numbers with unidentified origins. In addition, the Japanese government pushed through the domestic tourism campaign program in June 2020, with SNCDC backing, despite opposition from several prefectural governors and public opinion.[33, 34] Moreover, Miyawaki et al. (2021) found that domestic tourism campaign program participants had higher incidence rates of COVID-19 like symptoms than non-participants and concluded that the program increased COVID-19 cases in Japan.[35]

Turning to the planned Tokyo Olympic and Paralympic Games, as of June 22, 2021, the SNCDC has offered no detailed management strategies. Despite global scientific criticism and advice on preventing transmission, the Tokyo Olympic and Paralympic Games went off as scheduled between April and May 2021.[9, 10] Normally, scientific advisory bodies like the SNCDC provide management recommendations for preventing SARS-CoV-2 transmission at such events. There remains no valid explanation of why the SNCDC members backed the Japanese government in deviating from established global strategies for preventing SARS-CoV-2 transmission. Nevertheless, several Japanese newspapers reported that the government appointed experts to the SNCDC to advocate for the government[36, 37] and might also have prevented dissenting SNCDC members from publishing any conflicting views.

Nevertheless, apparent COI existed between those desiring to hold the Tokyo Olympic and Paralympic Games for economic and political purposes and public health experts’ concerns based on rigorous scientific evidence.[38] On June 18, 2021, 26 experts, including nine SNCDC members, voluntarily published their recommendations to the Japanese government and the International Olympic Committee on hosting the Tokyo Olympic and Paralympic Games, not as governmental advisory board members but as independent experts.[39] Our findings suggest that close relationships between the SNCDC members and the government, combined with the worst possible COI transparency policies, might reasonably explain this issue. At very least, we recommend that the Japanese government provide full disclosure of SNCDC members COI for the past three years and incorporate input from clinicians and subject matter experts with experience from different disciplines such as geneticists, data scientists, immunologists, statisticians, and infectious disease and public health experts with alternative views.

These same strategies already provide the basis for establishing trustworthy clinical practice guidelines widely used throughout Japan.[40, 41] Presently, the Japanese government must reconsider its priorities, economic growth based on the inadequate advice from muzzled experts or rigorous scientifically backed evidence governing sound public health policy.

This study has several limitations. First, due to the limited data availability, the time frame of disclosed pharmaceutical company payments did not overlap concisely with the time frame of advisory board member COI disclosure, as we have acknowledged in this study. Nevertheless, this study still provides insight into the magnitude of undeclared COI among the SNCDC members have with pharmaceutical companies and the Japanese government. Second, we assessed COI between the SNCDC members and the government-funded research grants on infectious diseases from the Ministry of Health, Labor, and Welfare. Although there may be other sources of research grants from the Japanese government, grants for infectious disease studies would most likely directly impact the opinions and suggestions of SNCDC members. Despite these limitations, to the best of our knowledge, this is the first study comprehensively assessing financial and non-financial COI among the Japanese government COVID-19 advisory board.

## Conclusions

We found that COVID-19 advisory board members had financial and non-financial COI with pharmaceutical companies and the Japanese government. Additionally, there were no rigorous management strategies for COI declaration and government disclosure to the public. This lack of proper COI management might unduly influence advisory board members’ recommendations. Therefore, the Japanese government must move quickly to ensure the independence of scientific advisory committees and implement more rigorous and transparent COI management strategies that include full declaration and disclosure to the public.

## Data Availability

All data are available in the Supplementary Materials

## Acknowledgment

The authors appreciate the Tansa for collecting payment data, Ms. Erika Yamashita for organizing the data, and Dr. Derek Hagman for professional language editing.

## Abbreviations

COI: conflicts of interest
COVID-19: coronavirus disease 2019
EMNCDCC: Expert Meeting on Novel Coronavirus Disease Control Committee
SARS-CoV-2: severe acute respiratory syndrome coronavirus 2
SNCDC: Subcommittee on Novel Coronavirus Disease Control
U.S.: United States

## Author contributions

We describe contributions to the paper using the CRediT taxonomy provided above.

Conceptualization: all authors;

Data curation: A.M.;

Formal analysis: H.M, A.M.;

Funding acquisition: A.O.;

Investigation: all authors; Methodology: all authors;

Project Administration: A.O., and T.T.;

Supervision: A.O., and T.T;

Visualization: H.M, and A.M;

Writing – Original Draft: all authors;

Writing – review & editing: A.M., T.T., and A.O.

All authors revised the manuscript critically for important intellectual content, gave final approval of the revision to be published, and agreed to be accountable for all the aspects of the study.

## Supporting information

**Supplemental Material 1. Five largest-paying pharmaceutical companies and their involvement in COVID-19**.

| Ranking | Company name | Personal<br>payments, US\$ | Total<br>payments, US\$ | Involvement in COVID-19 |
| --- | --- | --- | --- | --- |
| 1 | FujiFilm | 32,859 | 86,657 | Potential drug for treating COVID-19, favipiravir (Avigan), is in Phase 3 trial sponsored by the Ministry of Health, Labor, and Welfare. |
| 2 | Daiichi Sankyo | 15,414 | 62,968 | mRNA COVID-19 vaccine is currently in Phase 1/2 clinical trials<br>The manufacturer of the COVID-19 vaccine developed by AstraZeneca in Japan |
| 3 | Pfizer, | 52,882 | 61,871 | COVID-19 vaccines were the first approved in Japan |
| 4 | Sumitomo<br>Dainippon Pharma | 4,012 | 30,980 | None |
| 5 | Astellas Pharma | 25,131 | 29,591 | Providing potential compounds for the treatment of COVID-19 in response to a call from the Ministry of Health, Labor, and Welfare |

**Supplemental Material 2. Payment Data**

